# Highly multiplexed cytokine analysis of bronchoalveolar lavage and plasma reveals age-related dynamics and correlates of inflammation in children

**DOI:** 10.1101/2024.06.28.24309620

**Authors:** Shivanthan Shanthikumar, Liam Gubbels, Karen Davies, Hannah Walker, Anson Tsz Chun Wong, Eric Levi, Richard Saffery, Sarath Ranganathan, Melanie R. Neeland

## Abstract

Despite the central role of cytokines in mediating inflammation that underlies a range of childhood diseases, cytokine testing remains primarily limited to research settings and surrogate markers of inflammation are often used to inform clinical diagnostic and treatment decisions. There are currently no reference ranges available for cytokines in healthy children, either systemically (in blood) or at sites of disease (such as the lung). In our study, we aimed to develop an openly accessible reference of cytokines in the airways and blood of healthy children spanning 1 to 16 years of age. We examined how cytokine concentration changes during childhood and assessed whether a core set of cytokine markers could be used to indirectly evaluate the response of a broad spectrum of inflammatory analytes. To develop our reference, a total of 78 unique analytes were measured in cell-free bronchoalveolar lavage (BAL) and plasma from 78 children. We showed that age profoundly impacts soluble immune analyte concentration in both sample types and identified a highly correlative core set of 10 analytes in BAL and 11 analytes in plasma capable of indirectly evaluating the response of up to 34 inflammatory mediators. This study addresses an urgent need to develop reference ranges for cytokines in healthy children to aid in diagnosis of disease, to determine eligibility for, and to monitor the effects of, cytokine-targeted monoclonal antibody therapy.

## INTRODUCTION

While some established circulating blood markers, such as c-reactive protein (CRP), erythrocyte sedimentation rate (ESR) and ferritin, are used clinically to assess inflammation in children, cytokine testing is primarily limited to research settings (*1*). As a result, clinicians often use surrogate markers of cytokines to inform diagnostic and treatment decisions. For example, CRP level is often used as a surrogate of IL-6 and in patients with severe COVID-19 pneumonia and paediatric multisystem inflammatory syndrome temporarily associated with SARS-CoV-2 (PIMS-TS), CRP is used to inform treatment with tocilizumab (an anti-IL-6 monoclonal antibody) (*2, 3*). Other monoclonal antibodies such as those that target IL-4, IL-5, IL-13 and thymic stromal lymphopoietin (TSLP) are used for children with severe asthma, however surrogate markers such as blood eosinophil count or fractional exhaled nitric oxide are used to determine eligibility for these treatments (*4*). There is therefore a clear and urgent need to develop reference ranges for cytokines in healthy children to aid in diagnosis of disease, to determine eligibility for, and to monitor the effects of, immunomodulatory treatments (*5*).

The development of reference ranges for systemic cytokines will have wide applicability, as blood is currently used to assess for inflammation as well as for alterations in immune cell populations in a range of childhood immune-related conditions (*6–8*). For diseases that affect specific organs, reference values that reflect the local tissue are also needed to inform targeted treatment strategies. For example, respiratory diseases such as asthma and acute respiratory tract infections are major sources of morbidity and hospitalisation for children worldwide, yet treatment options are limited and often confer little to no clinical benefit (*9*). The development of disease modifying or curative treatment strategies in these children relies on a better understanding of immunity in the early healthy airway.

To address this, we aimed to develop an age-specific and extensive reference of cytokines and other soluble immune analytes in the airways and blood of children. We also aimed to investigate how immune analyte concentration changes during childhood and whether a specific core set of markers could be used to reflect inflammation both locally and systemically.

## RESULTS

### 1. Establishing a reference for immune analytes the lower airway and blood of children

We sampled bronchoalveolar lavage (BAL) from 24 children and blood from 66 children undergoing general anaesthesia at the Royal Children’s Hospital Melbourne (Figure 1A). The clinical indications for these procedures are provided in Table 1, with 92% (22/24) of the BAL cohort undergoing a procedure for investigation of stridor, and 70% (46/66) of the blood cohort undergoing a procedure for obstructive sleep apnoea (OSA). All children were clinically well at the time of sample collection. Medical history and information on respiratory status from parent questionnaires are also provided in Table 1. All BAL samples were assessed for pathogens as per standard clinical testing in the RCH clinical microbiology laboratory, which included standard culture for bacteria and fungi.

**Figure 1.**
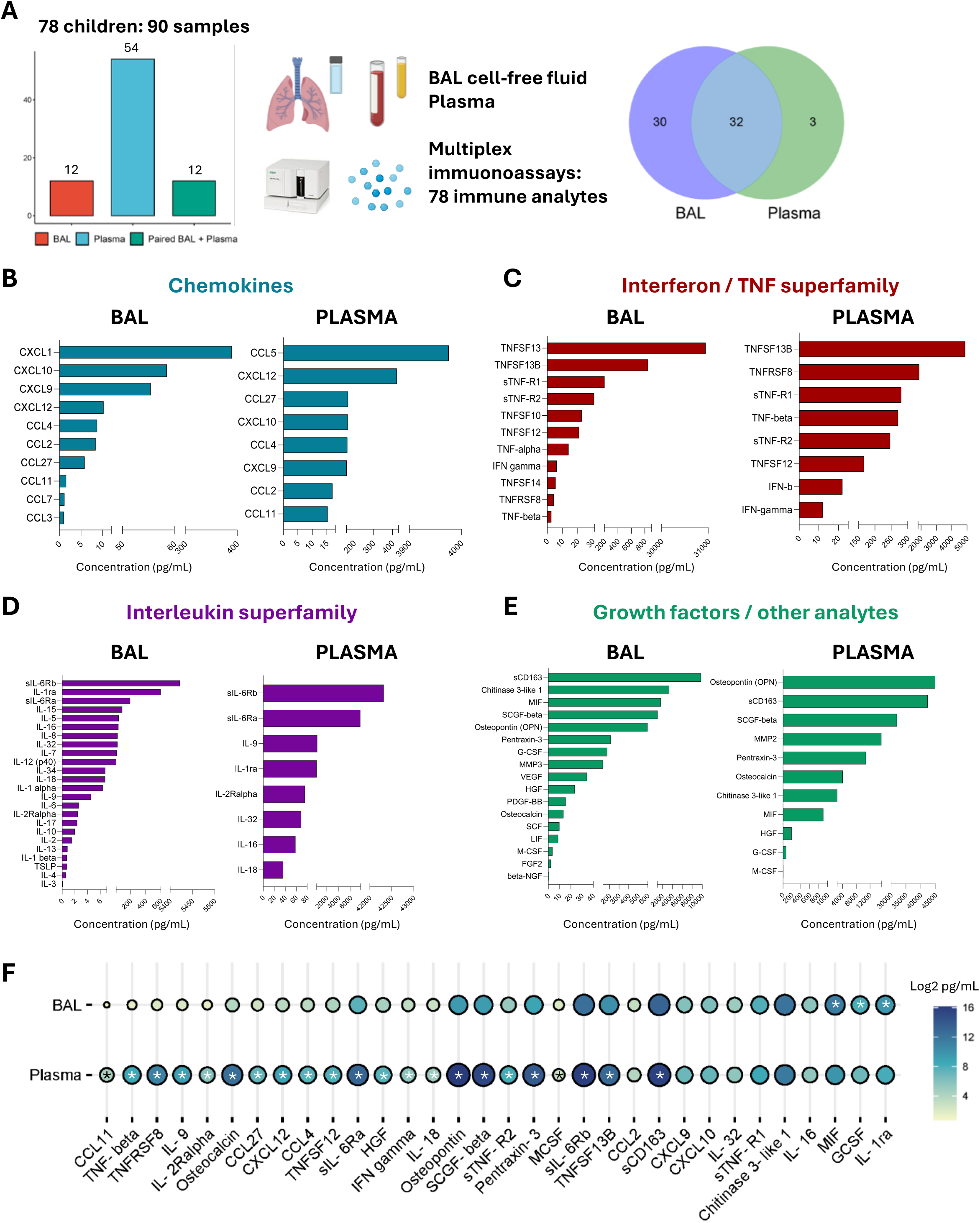
Landscape of soluble immune analytes in bronchoalveolar lavage (BAL) and plasma of healthy children. **(A)** Overview of the experimental design and a venn diagram depicting the overlap in the number of analytes detected in BAL vs plasma. Median concentration (pg/mL) of **(B)** chemokine family analytes, **(C)** interferon / TNF superfamily analytes, **(D)** interleukin superfamily analytes and **(E)** growth factors in BAL (n=24) and plasma (n=66). **(F)** Ballon plot depicting intra-individual comparison of analyte concentration between BAL and plasma for the 32 analytes detected across both samples in children who had both sample types collected (n=12). *Depicts significant difference by Wilcoxon signed-rank test and correction for the false discovery rate using the Benjamini Hochberg approach (FDR-p<0.05 was considered significant).

**Table 1.** Demographics and clinical characteristics of the study cohort.

|  | **BAL cohort** | **Plasma cohort** |
| --- | --- | --- |
| **Total number** | 24 | 66 |
| **Age: years, median (range)** | 3.31 (0.48-12.36) | 6.3 (0.48-16.27) |
| **Sex: male, n (%)** | 19 (79.2) | 40 (60.6) |
| **Reason for procedure, n (%)** |  |  |
| -OSA | 1 (4.2) | 46 (69.7) |
| -stridor | 22 (91.6) | 10 (15.15) |
| -other (hearing concerns,<br>nasal obstruction, tonsillitis) | 1 (4.2) | 10 (15.15) |
| **Familial atopy, n (%)** | 7 (29.2) | 37 (56.1) |
| **Personal atopy, n (%)** | 4 (16.2) | 25 (37.8) |
| **Parent report of doctor-**<br>**diagnosed asthma, n (%)** | 1 (4.2) | 18 (27.2) |
| **Any smoke exposure, n (%)** | 3 (12.5) | 16 (24.2) |
| **Antibiotic use past 12**<br>**months, n (%)** | 9 (37.5) | 22 (33.3) |
| **Inhaled steroid use past 12**<br>**months, n (%)** | 7 (29.2) | 8 (12.1) |
| **Oral steroid use past 12**<br>**months, n (%)** | 7 (29.2) | 14 (21.2) |

Participants were aged between 0.5 and 16 years, and we performed age stratification across infancy (0-2 years), preschool period (3-5 years), childhood (6-10 years), and adolescence (11-16 years). Of the 24 children with BAL samples, 12 had paired blood samples permitting a cross-tissue comparison of analyte levels across the two locations. A total of 78 unique analytes were measured in both the cell-free BAL and blood (plasma) samples using the 48-plex Bio-Plex Pro™ Human Cytokine Screening panel and the 37-plex Bio-Plex Pro™ Human Inflammation panel. All data are openly available for reuse in the data repository.

### 2. Landscape of immune analytes in BAL and plasma

Of the 78 analytes measured, 32 were detected in both samples, 30 were only detectable in BAL, 3 were only detectable in plasma, and 13 were not detectable in either sample type (Figure 1A, Supplemental Table 1). The cytokines that were undetectable in plasma in our study were also undetectable in another study using the same multiplex panel in plasma of a smaller number of healthy subjects (*10*). To understand and compare the landscape of soluble immune factors in BAL and plasma, we grouped analytes based on their functional properties.

This showed that while the concentrations of analytes differ between the blood and lung, the patterns of relative expression are similar between the two sample types, with CXCL10 and CXCL12 among the most abundant chemokines (Figure 1B), TNFSF13B, sTNF-R1, and sTNF-R2 the most abundant of the TNF superfamily (Figure 1C), sIL-6Ra, sIL-6Rb, and IL-1Ra the most abundant of the interleukin superfamily (Figure 1D), and inflammatory markers sCD163, SCGF-beta and Osteopontin also highly expressed in both sample types (Figure 1E).

There were 12 children in our cohort with paired BAL and plasma samples, enabling intra-individual analysis of analytes across both tissues (Figure 1F). This revealed that there were 22 analytes significantly more abundant in plasma compared to BAL (all FDR-p<0.01, Figure 1F). There were 3 analytes (IL-1Ra, G-CSF, MIF) significantly more abundant in BAL compared to plasma (all FDR-p<0.01, Figure 1F).

To help develop age-specific references for immune analytes in BAL and plasma, the median and range of each analyte are reported by clinically relevant age groups (infancy, preschool, childhood, adolescence) and presented in Tables 2-3.

**Table 2.**
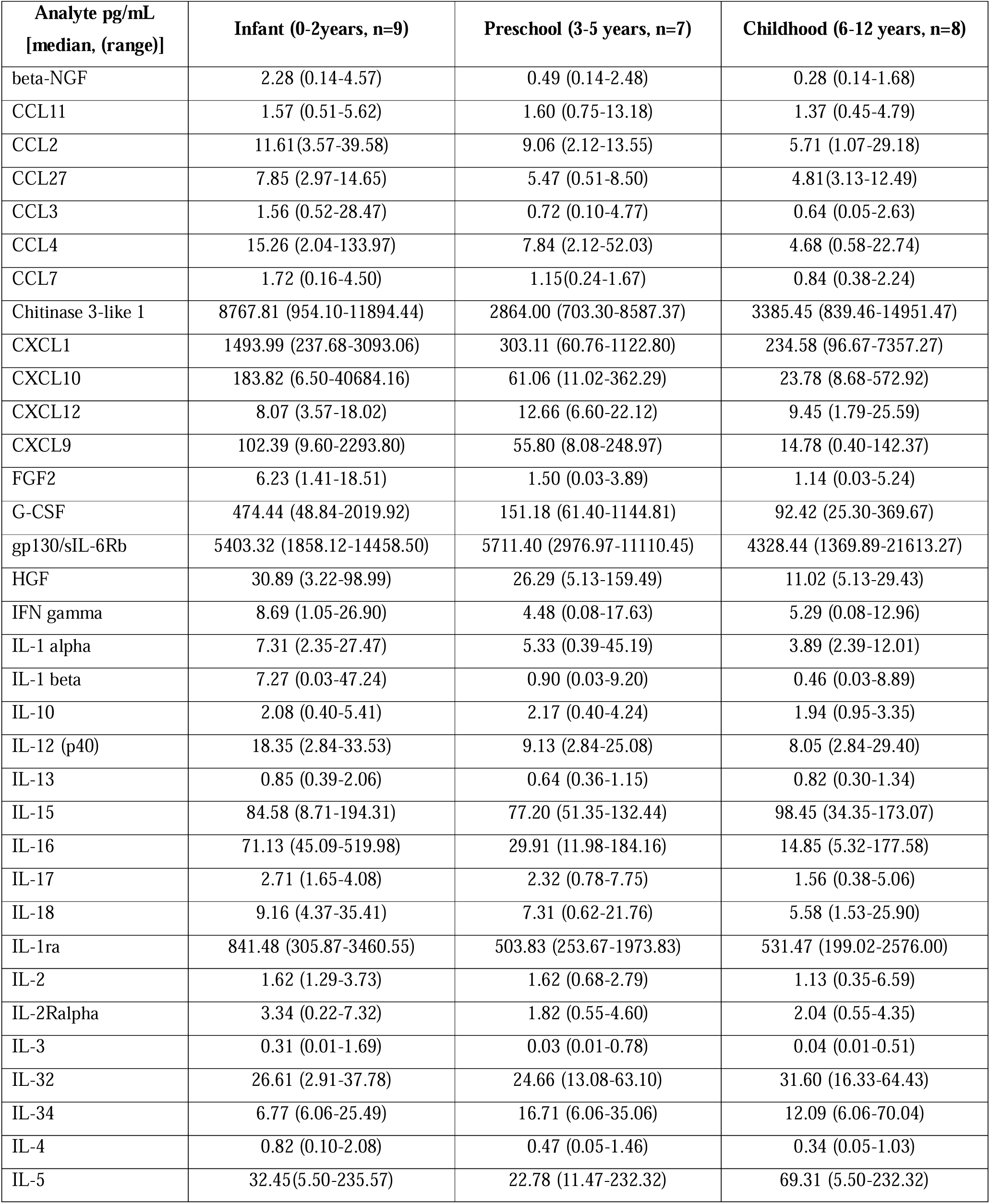
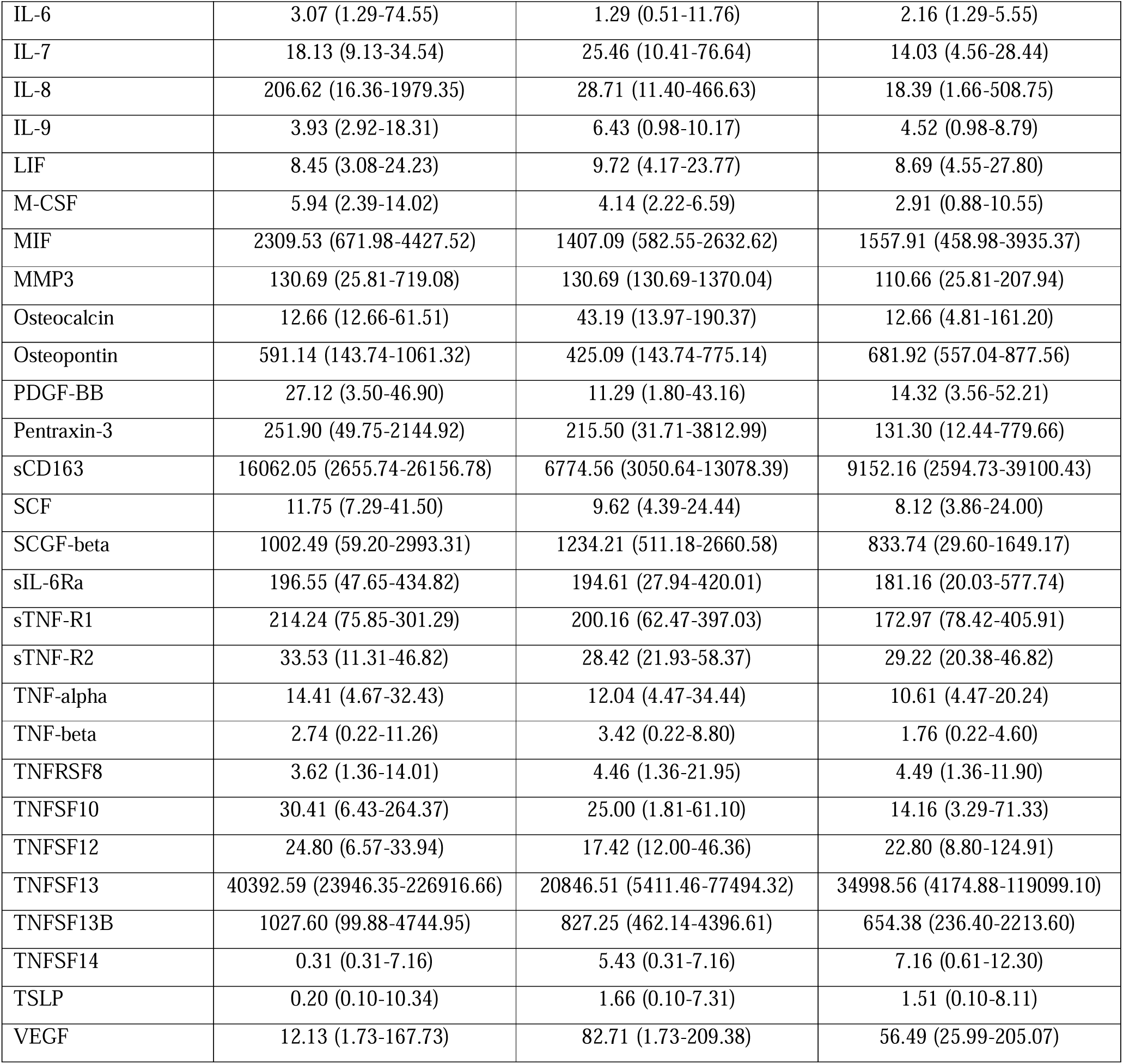
Immune analyte concentration in BAL cell-free fluid from children aged 0-12 years.

| Analyte pg/mL<br>[median, (range)] | Infant (0-2years, n=9) | Preschool (3-5 years, n=7) | Childhood (6-12 years, n=8) |
| --- | --- | --- | --- |
| beta-NGF | 2.28 (0.14-4.57) | 0.49 (0.14-2.48) | 0.28 (0.14-1.68) |
| CCL11 | 1.57 (0.51-5.62) | 1.60 (0.75-13.18) | 1.37 (0.45-4.79) |
| CCL2 | 11.61(3.57-39.58) | 9.06 (2.12-13.55) | 5.71 (1.07-29.18) |
| CCL27 | 7.85 (2.97-14.65) | 5.47 (0.51-8.50) | 4.81(3.13-12.49) |
| CCL3 | 1.56 (0.52-28.47) | 0.72 (0.10-4.77) | 0.64 (0.05-2.63) |
| CCL4 | 15.26 (2.04-133.97) | 7.84 (2.12-52.03) | 4.68 (0.58-22.74) |
| CCL7 | 1.72 (0.16-4.50) | 1.15(0.24-1.67) | 0.84 (0.38-2.24) |
| Chitinase 3-like 1 | 8767.81 (954.10-11894.44) | 2864.00 (703.30-8587.37) | 3385.45 (839.46-14951.47) |
| CXCL1 | 1493.99 (237.68-3093.06) | 303.11 (60.76-1122.80) | 234.58 (96.67-7357.27) |
| CXCL10 | 183.82 (6.50-40684.16) | 61.06 (11.02-362.29) | 23.78 (8.68-572.92) |
| CXCL12 | 8.07 (3.57-18.02) | 12.66 (6.60-22.12) | 9.45 (1.79-25.59) |
| CXCL9 | 102.39 (9.60-2293.80) | 55.80 (8.08-248.97) | 14.78 (0.40-142.37) |
| FGF2 | 6.23 (1.41-18.51) | 1.50 (0.03-3.89) | 1.14 (0.03-5.24) |
| G-CSF | 474.44 (48.84-2019.92) | 151.18 (61.40-1144.81) | 92.42 (25.30-369.67) |
| gp130/sIL-6Rb | 5403.32 (1858.12-14458.50) | 5711.40 (2976.97-11110.45) | 4328.44 (1369.89-21613.27) |
| HGF | 30.89 (3.22-98.99) | 26.29 (5.13-159.49) | 11.02 (5.13-29.43) |
| IFN gamma | 8.69 (1.05-26.90) | 4.48 (0.08-17.63) | 5.29 (0.08-12.96) |
| IL-1 alpha | 7.31 (2.35-27.47) | 5.33 (0.39-45.19) | 3.89 (2.39-12.01) |
| IL-1 beta | 7.27 (0.03-47.24) | 0.90 (0.03-9.20) | 0.46 (0.03-8.89) |
| IL-10 | 2.08 (0.40-5.41) | 2.17 (0.40-4.24) | 1.94 (0.95-3.35) |
| IL-12 (p40) | 18.35 (2.84-33.53) | 9.13 (2.84-25.08) | 8.05 (2.84-29.40) |
| IL-13 | 0.85 (0.39-2.06) | 0.64 (0.36-1.15) | 0.82 (0.30-1.34) |
| IL-15 | 84.58 (8.71-194.31) | 77.20 (51.35-132.44) | 98.45 (34.35-173.07) |
| IL-16 | 71.13 (45.09-519.98) | 29.91 (11.98-184.16) | 14.85 (5.32-177.58) |
| IL-17 | 2.71 (1.65-4.08) | 2.32 (0.78-7.75) | 1.56 (0.38-5.06) |
| IL-18 | 9.16 (4.37-35.41) | 7.31 (0.62-21.76) | 5.58 (1.53-25.90) |
| IL-1ra | 841.48 (305.87-3460.55) | 503.83 (253.67-1973.83) | 531.47 (199.02-2576.00) |
| IL-2 | 1.62 (1.29-3.73) | 1.62 (0.68-2.79) | 1.13 (0.35-6.59) |
| IL-2Ralpha | 3.34 (0.22-7.32) | 1.82 (0.55-4.60) | 2.04 (0.55-4.35) |
| IL-3 | 0.31 (0.01-1.69) | 0.03 (0.01-0.78) | 0.04 (0.01-0.51) |
| IL-32 | 26.61 (2.91-37.78) | 24.66 (13.08-63.10) | 31.60 (16.33-64.43) |
| IL-34 | 6.77 (6.06-25.49) | 16.71 (6.06-35.06) | 12.09 (6.06-70.04) |
| IL-4 | 0.82 (0.10-2.08) | 0.47 (0.05-1.46) | 0.34 (0.05-1.03) |
| IL-5 | 32.45(5.50-235.57) | 22.78 (11.47-232.32) | 69.31 (5.50-232.32) |
| IL-6 | 3.07 (1.29-74.55) | 1.29 (0.51-11.76) | 2.16 (1.29-5.55) |
| IL-7 | 18.13 (9.13-34.54) | 25.46 (10.41-76.64) | 14.03 (4.56-28.44) |
| IL-8 | 206.62 (16.36-1979.35) | 28.71 (11.40-466.63) | 18.39 (1.66-508.75) |
| IL-9 | 3.93 (2.92-18.31) | 6.43 (0.98-10.17) | 4.52 (0.98-8.79) |
| LIF | 8.45 (3.08-24.23) | 9.72 (4.17-23.77) | 8.69 (4.55-27.80) |
| M-CSF | 5.94 (2.39-14.02) | 4.14 (2.22-6.59) | 2.91 (0.88-10.55) |
| MIF | 2309.53 (671.98-4427.52) | 1407.09 (582.55-2632.62) | 1557.91 (458.98-3935.37) |
| MMP3 | 130.69 (25.81-719.08) | 130.69 (130.69-1370.04) | 110.66 (25.81-207.94) |
| Osteocalcin | 12.66 (12.66-61.51) | 43.19 (13.97-190.37) | 12.66 (4.81-161.20) |
| Osteopontin | 591.14 (143.74-1061.32) | 425.09 (143.74-775.14) | 681.92 (557.04-877.56) |
| PDGF-BB | 27.12 (3.50-46.90) | 11.29 (1.80-43.16) | 14.32 (3.56-52.21) |
| Pentraxin-3 | 251.90 (49.75-2144.92) | 215.50 (31.71-3812.99) | 131.30 (12.44-779.66) |
| sCD163 | 16062.05 (2655.74-26156.78) | 6774.56 (3050.64-13078.39) | 9152.16 (2594.73-39100.43) |
| SCF | 11.75 (7.29-41.50) | 9.62 (4.39-24.44) | 8.12 (3.86-24.00) |
| SCGF-beta | 1002.49 (59.20-2993.31) | 1234.21 (511.18-2660.58) | 833.74 (29.60-1649.17) |
| sIL-6Ra | 196.55 (47.65-434.82) | 194.61 (27.94-420.01) | 181.16 (20.03-577.74) |
| sTNF-R1 | 214.24 (75.85-301.29) | 200.16 (62.47-397.03) | 172.97 (78.42-405.91) |
| sTNF-R2 | 33.53 (11.31-46.82) | 28.42 (21.93-58.37) | 29.22 (20.38-46.82) |
| TNF-alpha | 14.41 (4.67-32.43) | 12.04 (4.47-34.44) | 10.61 (4.47-20.24) |
| TNF-beta | 2.74 (0.22-11.26) | 3.42 (0.22-8.80) | 1.76 (0.22-4.60) |
| TNFRSF8 | 3.62 (1.36-14.01) | 4.46 (1.36-21.95) | 4.49 (1.36-11.90) |
| TNFSF10 | 30.41 (6.43-264.37) | 25.00 (1.81-61.10) | 14.16 (3.29-71.33) |
| TNFSF12 | 24.80 (6.57-33.94) | 17.42 (12.00-46.36) | 22.80 (8.80-124.91) |
| TNFSF13 | 40392.59 (23946.35-226916.66) | 20846.51 (5411.46-77494.32) | 34998.56 (4174.88-119099.10) |
| TNFSF13B | 1027.60 (99.88-4744.95) | 827.25 (462.14-4396.61) | 654.38 (236.40-2213.60) |
| TNFSF14 | 0.31 (0.31-7.16) | 5.43 (0.31-7.16) | 7.16 (0.61-12.30) |
| TSLP | 0.20 (0.10-10.34) | 1.66 (0.10-7.31) | 1.51 (0.10-8.11) |
| VEGF | 12.13 (1.73-167.73) | 82.71 (1.73-209.38) | 56.49 (25.99-205.07) |

**Table 3.** Immune analyte concentration in plasma from children aged 0-16 years.

| Analyte pg/mL<br>[median, (range)] | Infant<br>(0-2years, n=10) | Preschool<br>(3-5 years, n=21) | Childhood<br>(6-10 years, n=22) | Adolescence<br>(11-16 years, n=13) |
| --- | --- | --- | --- | --- |
| CCL11 | 23.18 (9.58-84.99) | 19.38 (1.10-45.87) | 13.15 (5.50-57.09) | 9.98 (1.10-36.13) |
| CCL2 | 23.49 (5.58-44.37) | 17.15 (3.39-51.76) | 15.99 (1.74-30.91) | 13.66 (1.74-30.91) |
| CCL27 | 198.40 (117.10-293.63) | 173.03 (100.03-399.09) | 162.13 (104.22-331.70) | 145.79 (72.49-320.53) |
| CCL4 | 172.80 (119.46-214.33) | 192.20 (97.87-234.36) | 153.79 (112.18-263.28) | 144.31 (101.71-201.84) |
| CCL5 | 4177.71<br>(2713.73-8015.05) | 4202.67<br>(1033.84-8204.98) | 3983.05<br>(2143.14-7582.60) | 2829.16<br>(1048.29-6880.26) |
| Chitinase 3-like 1 | 2596.10<br>(1552.31-7024.70) | 2516.86<br>(733.91-3752.57) | 2373.99<br>(1384.95-4157.89) | 2742.96<br>(619.10-8551.33) |
| CXCL10 | 212.05 (96.42-543.98) | 228.10 (26.77-820.49) | 122.72 (47.46-380.63) | 77.61 (47.46-294.65) |
| CXCL12 | 408.73 (160.90-820.30) | 451.94 (195.36-1200.98) | 421.68 (233.94-613.11) | 398.93 (257.78-749.61) |
| CXCL9 | 430.80 (93.85-1071.94) | 269.87 (50.77-2985.30) | 155.22 (58.26-678.40) | 88.07 (41.55-219.88) |
| G-CSF | 116.57 (24.76-152.22) | 103.97 (16.80-461.44) | 54.95 (16.80-229.13) | 44.63 (16.80-103.97) |
| gp130/sIL-6Rb | 43169.60<br>(31111.97-66378.76) | 42195.48<br>(16316.25-57797.16) | 42237.24<br>(26541.97-88045.70) | 42245.00<br>(27559.36-98784.15) |
| HGF | 212.31 (160.90-484.38) | 191.34 (77.71-393.44) | 182.28 (116.16-498.00) | 209.38 (97.01-302.33) |
| IFN-b | 27.89 (17.93-47.31) | 23.09 (2.38-40.77) | 18.21 (11.02-44.00) | 20.47 (6.14-28.63) |
| IFN-y | 19.96 (4.86-107.49) | 15.45 (2.43-83.70) | 12.02 (2.43-39.02) | 4.86 (2.43-19.46) |
| IL-16 | 42.84 (31.28-110.58) | 61.34 (21.87-293.72) | 69.82 (15.40-319.00) | 58.40 (24.99-161.52) |
| IL-18 | 61.82 (24.97-153.02) | 45.29 (7.76-215.64) | 26.99 (0.83-84.66) | 27.53 (9.58-56.16) |
| IL-1ra | 286.16 (173.27-543.33) | 294.71 (149.56-556.97) | 328.53 (119.09-503.00) | 303.63 (50.45-520.95) |
| IL-2Ralpha | 109.62 (85.51-336.83) | 94.91 (40.24-192.19) | 68.78 (25.12-126.38) | 49.05 (17.79-95.79) |
| IL-32 | 87.05 (35.12-141.98) | 60.58 (17.77-129.91) | 64.98 (12.41-117.79) | 56.28 (5.31-100.00) |
| IL-9 | 445.69 (327.58-578.53) | 480.18 (241.67-561.13) | 385.53 (259.31-698.90) | 388.27 (288.37-624.03) |
| M-CSF | 13.90 (7.50-35.84) | 10.57 (1.60-36.80) | 7.62 (1.60-23.62) | 6.90 (1.60-16.63) |
| MIF | 726.01 (428.10-1290.45) | 939.32 (205.67-1809.94) | 876.66 (385.50-2257.42) | 1049.03 (207.09-1700.9) |
| MMP2 | 17340.48<br>(8729.47-36734.68) | 16119.55<br>(6708.58-27039.07) | 12801.61<br>(5844.31-34301.90) | 16114.69<br>(1750.16-34558.64) |
| Osteocalcin | 4043.32<br>(2792.44-5887.91) | 3970.84<br>(1702.10-8338.08) | 4057.51<br>(1304.42-7272.78) | 4106.13<br>(1332.68-8915.56) |
| Osteopontin | 47020.08<br>(33621.91-99030.07) | 41887.55<br>(16300.23-70515.61) | 44788.74<br>(24948.42-81080.82) | 46935.05<br>(20099.78-111027.38) |
| Pentraxin-3 | 12686.39<br>(6965.05-26118.35) | 9822.07<br>(1796.25-23211.50) | 11127.18<br>(4714.23-27852.32) | 8256.14<br>(1383.34-31842.04) |
| sCD163 | 56866.33<br>(32788.18-183229.97) | 43004.83<br>(12842.22-142352.08) | 37352.46<br>(19757.93-102636.90) | 41664.56<br>(17482.61-93158.19) |
| SCGF-beta | 38390.39<br>(28917.70-57447.48) | 35604.97<br>(16248.82-47686.49) | 28505.78<br>(13325.99-46096.23) | 32903.42<br>(12856.50-54756.44) |
| sIL-6Ra | 6605.90<br>(5614.23-15726.64) | 7915.80<br>(1820.17-14690.32) | 6852.21<br>(3829.03-12435.15) | 8314.40<br>(3028.12-10335.88) |
| sTNF-R1 | 448.58 (250.07-897.30) | 264.12 (116.94-562.76) | 289.89 (133.21-612.06) | 267.18 (43.34-771.93) |
| sTNF-R2 | 299.79 (243.75-461.46) | 262.02 (108.98-368.96) | 231.30 (179.17-357.47) | 237.42 (139.83-370.86) |
| TNF-beta | 312.84 (211.90-385.36) | 291.32 (172.50-406.08) | 255.43 (164.04-456.77) | 231.05 (166.32-327.68) |
| TNFRSF8 | 2982.19<br>(1900.97-8806.88) | 2800.79<br>(505.38-7161.09) | 1727.14<br>(681.48-2687.69) | 1019.36<br>(449.76-1790.75) |
| TNFSF12 | 184.89 (79.94-271.97) | 166.51 (70.70-241.90) | 169.01 (103.07-274.80) | 144.77 (56.60-323.16) |
| TNFSF13B | 5291.65<br>(3620.37-9854.53) | 4879.07<br>(1975.04-7382.17) | 5058.65<br>(2458.21-9941.11) | 4794.56<br>(3013.45-11110.11) |

### 3. Immune analyte dynamics over the first 16 years of life

Principal components analysis (PCA) of BAL analyte data was conducted including age, sex, and BAL pathogen status as demographic variables. This showed that sex and pathogen status did not significantly contribute to variation in our dataset, however age trended towards correlation with the first dimension PC-1 (R=0.238, p=0.13) which explained 36.96% of the variation (Figure 2A). To explore this further, we performed two-tailed spearman correlation analyses of the BAL analyte data which showed 21 analytes were significantly associated with age (raw p<0.05, Figure 2B). The BAL concentration of TNFSF14 and VEGF increased with age, while the concentration of IL-16, FGF2, beta-NGF, IL-4, IL-8, HGF, CXCL1, GCSF, CXCL9, CXCL10, CCL3, IL-2Ralpha, CCL7, CCL27, CCL2, CCL4, SCF, MCSF, and IL-1beta decreased with age. Protein-protein interaction (PPI) and enrichment analysis using STRING revealed that these 21 analytes were highly connected, showing enrichment of pathways associated with chemokine expression and binding, as well as PI3K-Akt signaling (Figure 2C). Of these 21 analytes, only TNFSF14, IL-16 and FGF-2 were significantly associated with age after correction for the false discovery rate (all FDR-p<0.05). Individual dot plots depicting correlations for each of these 21 analytes are presented in Supplementary Figure 1.

**Figure 2.**
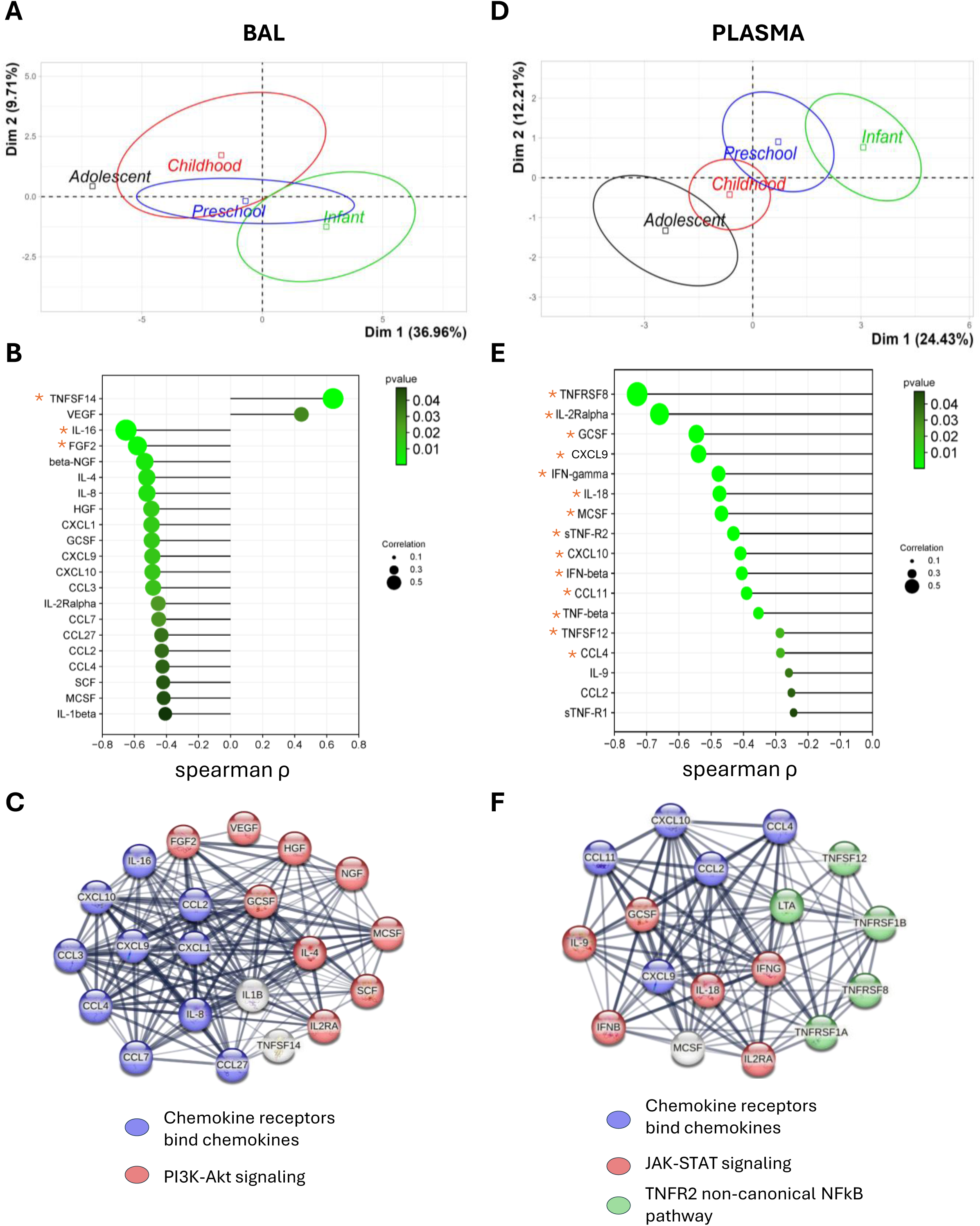
Immune analyte dynamics over the first 16 years of life. **(A)** Principal components analysis (PCA) of BAL immune analyte data highlighting age as a significant contributor to the first dimension. **(B)** Spearman coefficient and p-values for BAL analytes that were significantly associated with age. **\***Indicates analytes that were significant after correction for the false discovery rate (FDR). **(C)** Protein-Protein Interaction (PPI) analysis using STRING reveals significant interactions between BAL analytes associated with age (the strength of the line indicates the confidence of the interaction) and shows enrichment of pathways associated with chemokine receptor binding and PI3K-Akt signaling. **(D)** PCA of plasma immune analyte data highlighting age as a significant contributor to the first dimension. **(E)** Spearman coefficient and p-values for plasma analytes that were significantly associated with age. **\***Indicates analytes that were significant after FDR correction. **(F)** PPI analysis using STRING reveals significant interactions between plasma analytes associated with age and shows enrichment of pathways associated with chemokine receptor binding, JAK/STAT signaling and TNFR2 non-canonical NFκB activation. Correlation p-values were determined by two-tailed spearman test and p<0.05 was considered significant.

PCA of plasma analyte data was also conducted, including age and sex as demographic variables. This showed that sex did not significantly contribute to variation in the plasma dataset, however age was strongly correlated with the first two dimensions PC-1 (R=0.32, p=0.00002) and PC-2 (R =0.16, p=0.009) which together explained 36.6% of the variation (Figure 2D). Two-tailed spearman correlation analyses of the plasma analyte data showed 17 analytes were significantly and negatively correlated with age (raw p<0.05, Figure 2E). PPI and enrichment analysis using STRING revealed that these 17 analytes were highly connected with enrichment of three pathways: chemokine expression and binding, JAK/STAT signaling, and TNFR2 non-canonical NFκB activation (Figure 2F). Of these 17 analytes, 14 remained significantly associated with age after correction for the false discovery rate (all FDR-p<0.05). These were: TNFRSF8, IL-2Ralpha, GCSF, CXCL9, IFN-gamma, IL-18, MCSF, sTNF-R2, CXCL10, IFN-beta, CCL11, TNF-beta, TNFSF12, CCL4 (Figure 2E). Individual dot plots depicting correlations for all 17 analytes are presented in Supplementary Figure 2.

### 4. Sex and pathogen status of BAL do not largely modify immune analyte concentration in childhood

While sex and BAL pathogen status did not appear to contribute to variation by PCA, we wanted to confirm this by directly comparing analyte concentration in males and females for both the BAL and plasma data, as well as pathogen vs no pathogen detected for the BAL data. This revealed that BAL concentrations of IL-9 and IL-10 were slightly higher in males compared to females, and that BAL TNFRSF8 was slightly lower in females compared to males (Supplementary Figure 3A). Plasma concentrations of SCGF-beta, TNFSF13B and IFN-beta were higher in males compared to females (Supplementary Figure 3B). Of these six analytes, only plasma SCGF-beta was significantly different between males and females after correction for the false discovery rate (FDR-p=0.005) (Supplementary Figure 3B). There were no significant differences in BAL analyte concentration in samples with a pathogen detected compared to samples without a pathogen detected. A list of pathogens detected in our cohort is provided in Supplemental Table 2.

### 5. Discovery of a core set of highly correlated analytes to facilitate clinical application

While our comprehensive analysis has provided new insights into the soluble immune response in both BAL and plasma of children, not all analytes identified will have clinically accessible tests or a known clinical effect. To address this, we explored the feasibility of identifying a core group of analytes within each tissue that exhibit high correlation. This core set could then be measured in clinical settings to indirectly evaluate the response of other analytes. We also explored whether plasma analyte concentration correlated with BAL analyte concentration in individuals who had both sample types collected, with the goal of identifying markers that could act as proxies for the lung to reduce the need for invasive bronchoscopy for investigation of lung inflammation.

Intra-sample correlation analyses revealed robust correlation among analytes in BAL (Figure 3A), all with an FDR-p<0.001. Based on these highly correlated analytes, we identified a set of 10 core analytes that could estimate the concentrations of 34 other BAL analytes (Figure 3B). The core set of analytes comprises CCL2, Chitinase 3-like 1, FGF2, HGF, IL-17, IL-1alpha, IL-4, IL-5, sTNFR1, and TNFRSF8. Collectively, these analytes exhibit strong correlations with 34 other unique analytes: CCL27, CCL3, CCL4, CCL7, CXCL1, CXCL10, CXCL9, GCSF, IFN-gamma, IL-15, IL-16, IL-18, IL-1beta, IL-1ra, IL-2, IL-2Ralpha, IL-34, IL-6, IL-8, IL-9, LIF, MCSF, Pentraxin-3, sCD163, SCF, sIL-6Ra, sIL-6Rb, sTNFR2, TNF-alpha, TNF-beta, TNFSF10, TNFSF12, TNFSF14, and TNFSF13B.

**Figure 3.**
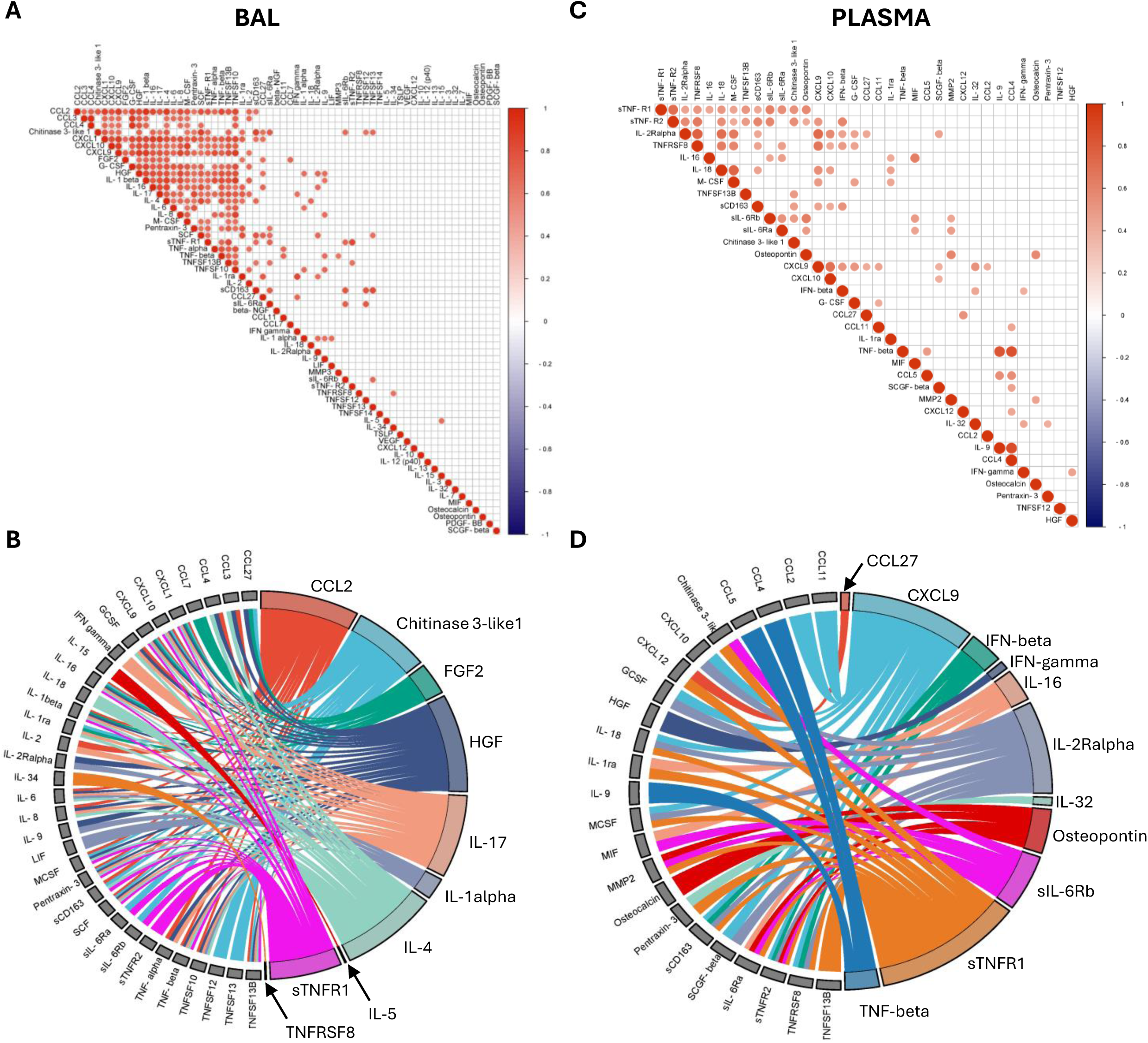
Discovery of a core set of highly correlated analytes. **(A)** Spearman correlation analyses of analytes within BAL. The red circles indicate a significant correlation after correction for the false discovery rate (all FDR-p<0.001). The strength of the colour indicates the strength of the correlation (spearman rho). **(B)** Chord plot depicting the BAL analytes with the highest number of correlations. This core set of analytes (right hand side) strongly correlate with 34 other unique analytes (left hand side). **(C)** Spearman correlation analyses of analytes within plasma. The red circles indicate a significant correlation after correction for the false discovery rate (all FDR-p<0.001). The strength of the colour indicates the strength of the correlation (spearman rho). **(D)** Chord plot depicting the plasma analytes with the highest number of correlations. This core set of analytes (right hand side) strongly correlate with 23 other unique analytes (left hand side).

Intra-sample correlation analyses also revealed high correlation among analytes in plasma (Figure 3C), all with an FDR-p<0.001. Based on these highly correlated analytes, we identified a set of 11 core analytes that could estimate the concentrations of 23 other plasma analytes (Figure 3D). The core set of analytes comprises CCL27, CXCL9, IFN-beta, IFN-gamma, IL-16, IL-2Ralpha, IL-32, Ostepontin, sIL-6Rb, sTNFR1, and TNF-beta. Collectively, these analytes exhibit strong correlations with 23 other unique analytes: CCL11, CCL2, CCL4, CCL5, Chitinase 3-like 1, CXCL10, CXCL12, GCSF, HGF, IL-18, IL-1ra, IL-9, MCSF, MIF, MMP2, Osteocalcin, Pentraxin-3, sCD163, SCGF-beta, sIL-6Ra, sTNFR2, TNFRSF8, TNFSF13B.

Cross-tissue correlation analyses of paired BAL and plasma samples revealed no significant correlations in analyte levels between the two tissue types in healthy children.

## DISCUSSION

We developed an extensive and openly accessible reference of soluble immune analytes in BAL and plasma of healthy children spanning 1 to 16 years of age. We showed that age profoundly impacts soluble immune analyte concentration in both sample types, highlighting the importance of developing age-specific reference datasets for immune responses in the airways and blood of children. We identified a highly correlative core set of 10 analytes in BAL and 11 analytes in plasma that could be measured to indirectly evaluate the response of a broad spectrum of inflammatory analytes, emphasising potential clinical feasibility of this work.

We showed that early life represents an increased state of inflammation compared to later childhood, with a significant number of analytes decreasing throughout childhood in both the airways and circulation. This reflects what has been observed previously in other studies of children and adults, where plasma analytes including CCL4, IL-18, IL-2Ralpha, CXCL9 and IFN-gamma were also shown to reduce with age (*10, 11*). Immune pathways associated with analytes that changed with age in our plasma data included chemokine receptor binding, JAK-STAT signaling and TNFR2 non-canonical NFκB activation. Increased JAK/STAT signaling is associated with several atopic conditions that are more common in children than adults (*12*). Furthermore, unregulated JAK/STAT signaling is a potential risk factor for chronic inflammation in the elderly (*13*). Non-canonical NFκB signaling regulates cell survival, maturation, and proliferation, as well as plays a key role in the development of immune tissues, highlighting the importance of these pathways in early life (*14*). We also recently showed that proportions of immune cell types associated with JAK/STAT and NFκB signaling, including conventional dendritic cells, monocytes, and B cells, also reduced with age in blood of children (*15*), providing a potential further link between these pathways and immune development in childhood.

To our knowledge, we are the first to report age-related changes in immune analyte concentration in BAL of healthy children. Like plasma, many analytes reduced with age in BAL, while two (TNFSF14 and VEGF) increased with age. Immune pathways associated with analytes that changed with age in our BAL data include chemokine receptor binding and PI3K-Akt signaling. The PI3K-AKIT signaling pathway has been shown to mediate growth and survival during early lung development in mouse models, and dysregulation of this pathway is associated with the development of lung fibrosis is later life (*16–19*). Understanding the normal trajectory of BAL cytokines as children age will allow clinical teams to identify aberrations that occur in different disease states, including the key pathways identified above, and facilitate the use of cytokines as diagnostic biomarkers.

We identified a highly correlative core set of 10 analytes in BAL and 11 analytes in plasma that could be measured to indirectly evaluate the response of a broad spectrum of inflammatory analytes. We developed these core sets of analytes as it is unlikely that all analytes investigated in our study will have a clinically available test and the ability to test a core set of markers as a surrogate for a broad array of immunological features is more clinically feasible. Using such an approach is not implausible, as CRP is currently used as a surrogate for IL-6 concentration, and a high level of CRP is used in clinical decision making around anti-IL6 medication (*2, 3*). Our core set of 11 plasma analytes allows evaluation of 34 total plasma markers, including IL-18, a key feature of childhood conditions including hemophagocytic lymphohistiocytosis (HLH) and juvenile idiopathic arthritis (*20, 21*), the TNF and IFN superfamilies which are biomarkers of several autoimmune and inflammatory diseases for which monoclonal antibody therapies are available (*22*), and osteopontin which has been proposed as a potential biomarker in paediatric cancer patients (*23*). Our core set of 10 BAL analytes allows evaluation of 44 total BAL markers, including IL-6 which is a key feature of multi system inflammatory syndrome following SARS-CoV-2 infection in children (*24*), IL-4, Pentraxin-3, and IL-5 which have all been associated with wheezing and asthma in children (*25, 26*), HGF and FGF2 which are key markers of pulmonary fibrosis seen in paediatric lung diseases of genetic (i.e. disorders of surfactant metabolism) or acquired (i.e. chemotherapy, hypersentivitiy pneumonitis, radiation) origin (*27, 28*), and the chemokine ligand family which are key players in lung diseases that involve inflammatory cell infiltration including paediatric cystic fibrosis (*29*).

Interestingly, we observed no significant correlation in analyte levels between plasma and BAL in children who had paired samples. This indicates that, in healthy children, systemic cytokine levels may not reflect those in the lung, although we acknowledge the smaller number of participants with matched samples for this analysis. These findings are similar to our previous work that showed no significant correlation in immune cell type proportions between healthy paediatric blood and airway samples (*15*). While our data suggest that circulating immune markers may not be good proxies for the lung immune response in healthy children, studies investigating respiratory conditions such as asthma, severe COVID-19 and cystic fibrosis have shown that blood markers, including IL-6 and eosinophils, positively correlate with those in BAL during disease (*29–31*).

We acknowledge our samples were collected as part of clinically indicated procedures, which means that not all samples can be considered ‘healthy’. All children were clinically well at the time of sample collection. The main indication in our plasma cohort was OSA, with 70% of samples obtained from children exhibiting or undergoing investigation for this indication. The effect of this underlying condition on our results obtained in plasma is unclear. A recent review revealed conflicting evidence of immune alterations in adult and paediatric OSA, with some studies reporting an inflammatory effect while others reported normal systemic immune responses in these patients (*32*). The main indication in our BAL cohort was stridor (>90% of the BAL cohort), a sign that affects the upper airway and not the lung and thus should not affect the BAL cytokine profile, and only 1 participant in the BAL cohort had a parent report of ever having asthma. Parent questionnaires revealed that the majority of children in this cohort had never been prescribed respiratory medications (inhaled or oral steroids), and of those prescribed, rates of medication use were similar to what is observed in the Australian community for children aged 0-15years (*33*). As general anaesthesia is required to collect BAL in children, creating control datasets from healthy children in the community is not feasible. Nevertheless, our cytokine reference can be used as control data for clinical samples collected from children with acute, rare, and chronic respiratory conditions.

In this study, we have shown age-related changes in cytokine concentration in both blood and BAL. We identified a core set of cytokines in each tissue that facilitates the clinical translation of our findings. Our work addresses a current knowledge gap regarding age-related trends in systemic and tissue specific cytokine levels that is important to address in the current era of cytokine targeted monoclonal antibody therapy for children in a range of diseases. Our findings will help improve our understanding of the role of inflammation in disease pathogenesis and develop improved biomarkers for disease diagnosis and therapeutic intervention.

## MATERIALS AND METHODS

### Study participants

This study took place at the Royal Children’s Hospital (RCH, Melbourne Australia) and involved analysis of 90 samples (24 BAL, 66 plasma) from 78 children (Table 1) aged between 1 and 16 years. Samples were collected at the time of clinically indicated procedures (bronchoscopy, tonsillectomy and/or adenoidectomy). Reasons for procedures, medical history, and respiratory status of the cohort are documented in Table 1. All children were clinically well at the time of the procedure.

### Ethics statement

The studies involving human participants were reviewed and approved by Royal Children’s Hospital Human Research Ethics Committee (HREC #25054 and # 88144). Written informed consent to participate in the studies was provided by the participants’ legal guardian/next of kin.

### Collection and processing of samples

For BAL samples, bronchoscopy and bronchoalveolar lavage were performed as previously described where indicated for investigation of stridor (*34*). This study used BAL from aliquots two and three from the right middle lobe. BAL was assessed for pathogens as per standard clinical testing in the RCH clinical microbiology laboratory, which included standard culture for bacteria and fungi. Up to 7mL of venous blood was collected in EDTA tubes at the time of the procedure. BAL samples were transferred on ice to the laboratory immediately upon collection. Blood samples were kept at room temperature and processed immediately after collection. BAL samples were centrifuged at 300xg for 7 minutes at 4°C and the cell-free BAL supernatant collected and stored at −80°C. EDTA blood was centrifuged at 700xg for 7 minutes at room temperature and the supernatant (plasma) stored at −80°C.

### Analyte quantification using Bio-Plex Pro panels

Cell-free BAL supernatant and plasma samples were thawed, and cytokines were measured using the 48-plex Bio-Plex Pro™ Human Cytokine Screening panel (Bio-Rad, California, USA) and the 37-plex Bio-Plex Pro™ Human Inflammation panel (Bio-Rad, California, USA) according to the manufacturer’s instructions. BAL supernatants were run undiluted and plasma samples were diluted 1:5 in phosphate-buffered saline (PBS) as per manufacturers recommendations. The full list of the 78 analytes measured is provided in Supplementary Table 1. Data were acquired on the Bio-Plex 200 system and data QC was done using the Bio-Plex Manager™ 6.1 software (Bio-Rad). For data QC, standard curve outliers were removed, and individual standards were adjusted to achieve a recovery rate of 100 ± 5% (observed concentration/expected concentration). A fit probability greater than 0.9 was achieved for each standard curve. The concentration values of each analyte control were then compared to the expected concentration range specified on the assay data sheet for the control lot. Analytes that were not detected in >50% of samples were excluded from further analysis. For each analyte included in the analysis, samples that fell below the detection range were arbitrarily reported as half the lower limit of detection of the assay as previously described for this assay (*35*). Samples were run over 5 plates for the 48-plex panel, and samples were run over 3 plates for the 37-plex panel. Plate (batch) was included as a variable in the analysis, detailed below.

### Data analysis

To identify sources of variation in our data, principal components analysis (PCA) was conducted on log transformed analyte data using the FactoMineR package, including age, sex, plate (batch), and respiratory pathogen status (for BAL data only) as variables. For respiratory pathogen status of BAL samples, this included ‘yes’ or ‘no’ for bacteria and/or fungi detected on culture at the RCH microbiology laboratory (there were no fungi detected in this cohort, see supplement for bacteria detected). As batch contributed significantly to the variation in both the BAL and plasma data (Supplementary Figure 4), we used the ComBat package to adjust for this batch effect. PCA analysis of ComBat-corrected data revealed that batch was effectively adjusted for, and the contribution of other factors including age, sex, and respiratory pathogen status remained unchanged (Supplementary Figure 4). Batch corrected values were used for all analysis presented herein however both uncorrected and batch corrected values are provided in the data repository. For age-specific immune analyte reference ranges (Tables 2-3), the median concentration (pg/mL) was calculated for each immune analyte within each age group (0-2years, 3-5 years, 6-10 years, 11-16 years) and the ranges also reported. There were 12 children in our cohort with paired BAL and plasma samples, enabling intra-individual analysis of analytes across both tissues. Wilcoxon signed-rank tests were used to compare the concentration of analytes between BAL and plasma in children who had both sample types collected (Figure 1F). FDR-corrected p-values are reported and FDR-p<0.05 was considered significant. FDR adjustment was done using the Benjamini and Hochberg approach (*36*). Age correlations were determined using two-tailed spearman tests (Figure 4B-E). Both raw (p<0.05) and FDR-corrected (FDR-p<0.05) p-values are reported. Analytes that were significantly correlated with age underwent protein-protein interactions (PPI) and pathway analysis using STRING (version 12) (*37*) (Figure 4C-F). Mann-Whitney U-tests were used to compare the concentration of analytes between males and females (Supplementary Figure 3, raw p-values reported, p<0.05 was considered significant) and BAL samples with a pathogen vs no pathogen detected (there were no significant differences). For intra sample type correlations (Figure 3), this was done using two-tailed spearman tests and FDR-corrected p-values are reported. To ensure the reliability of our findings, we present only analytes with FDR-p<0.001 for this analysis, which were used to generate the ‘core set’ of highly correlative analytes.

## Supporting information

Data repository

## Data Availability

All data produced in the present work are contained in the manuscript

## ACKNOWLEDGEMENTS

This work was funded by Chan Zuckerberg Initiative Single-Cell Biology Grants (2021-237883, 2020-217531). We thank the children and parents who participated in our studies, without whom this work would not exist.

