## Supplemental file for "Highly multiplexed cytokine analysis of bronchoalveolar lavage and plasma reveals age-related dynamics and correlates of inflammation in children"

**Supplementary Table 1.** List of analytes measured across the two BioPlex kits. Shaded boxes indicate the analyte was detectable in that sample type. There were 13 analytes not detectable in either BAL or plasma (no shading).

| Analyte | BAL | PLASMA | Analyte | BAL | PLASMA | Analyte | BAL | PLASMA |
|---|---|---|---|---|---|---|---|---|
| BAFF/TNFSF13B | | | sIL-6Ra | | | LIF | | |
| CTACK/CCL27 | | | sTNF-R1 | | | LIGHT/TNFSF14 | | |
| Eotaxin/CCL11 | | | sTNF-R2 | | | MCP-3/CCL7 | | |
| G-CSF | | | TNF-beta | | | MMP3 | | |
| gp130/sIL-6Rb | | | TWEAK/TNFSF12 | | | PDGF-BB | | |
| HGF | | | Chitinase 3-like 1 | | | SCF | | |
| IFN gamma | | | MIP-1alpha/CCL3 | | | TNF-alpha | | |
| IL-16 | | | APRIL/TNFSF13 | | | TRAIL/TNFSF10 | | |
| IL-18 | | | Basic FGF/FGF2 | | | TSLP | | |
| IL-1ra | | | beta-NGF | | | VEGF | | |
| IL-2Ralpha | | | GRO-alpha/CXCL1 | | | IFN-b | | |
| IL-32 | | | IL-1 alpha | | | MMP2 | | |
| IL-9 | | | IL-1 beta | | | RANTES/CCL5 | | |
| IP-10/CXCL10 | | | IL-10 | | | GM-CSF | | |
| MCP-1 /CCL2 | | | IL-12 (p40) | | | IFN-alpha 2 | | |
| M-CSF | | | IL-13 | | | IL-11 | | |
| MIF | | | IL-15 | | | IL-12(p70) | | |
| MIG/CXCL9 | | | IL-17 | | | IL-19 | | |
| MIP-1beta/CCL4 | | | IL-2 | | | IL-20 | | |
| Osteocalcin | | | IL-3 | | | IL-22 | | |
| Osteopontin (OPN) | | | IL-34 | | | IL-26 | | |
| Pentraxin-3 | | | IL-4 | | | IL-27 (p28) | | |
| sCD163 | | | IL-5 | | | IL-28A/IFN-lambda2 | | |
| sCD30/TNFRSF8 | | | IL-6 | | | IL-29/IFN-lambda1 | | |
| SCGF-beta | | | IL-7 | | | IL-35 | | |
| SDF-1alpha/CXCL12 | | | IL-8 | | | MMP1 | | |

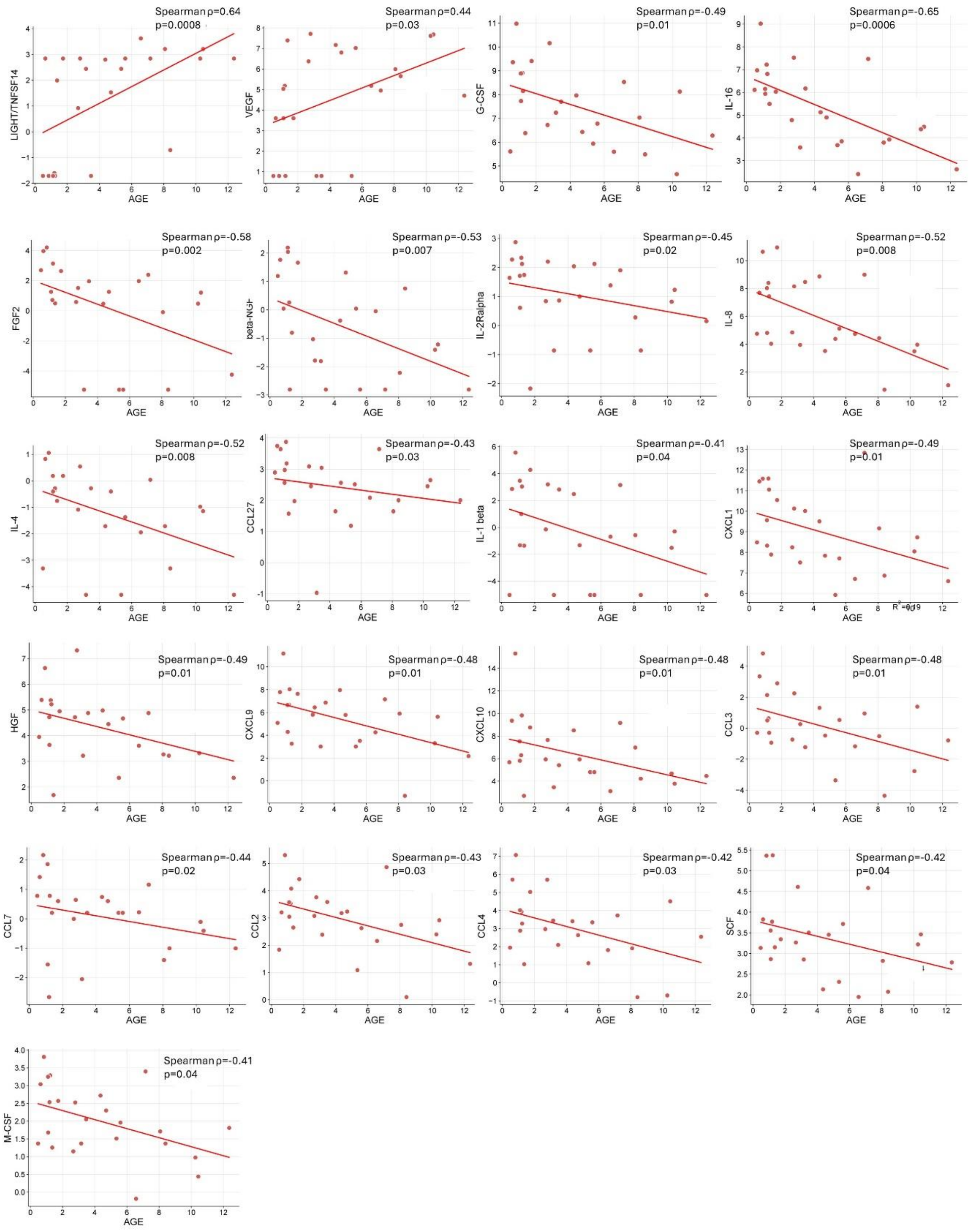


**Supplementary Figure 1.** Spearman correlation plots for each significant analyte in BAL. The plots depict age in years on the x-axis, and the concentration (log2 pg/mL) of each analyte on the y-axis. Spearman coefficient (ρ) and raw p-values are shown, n=24.

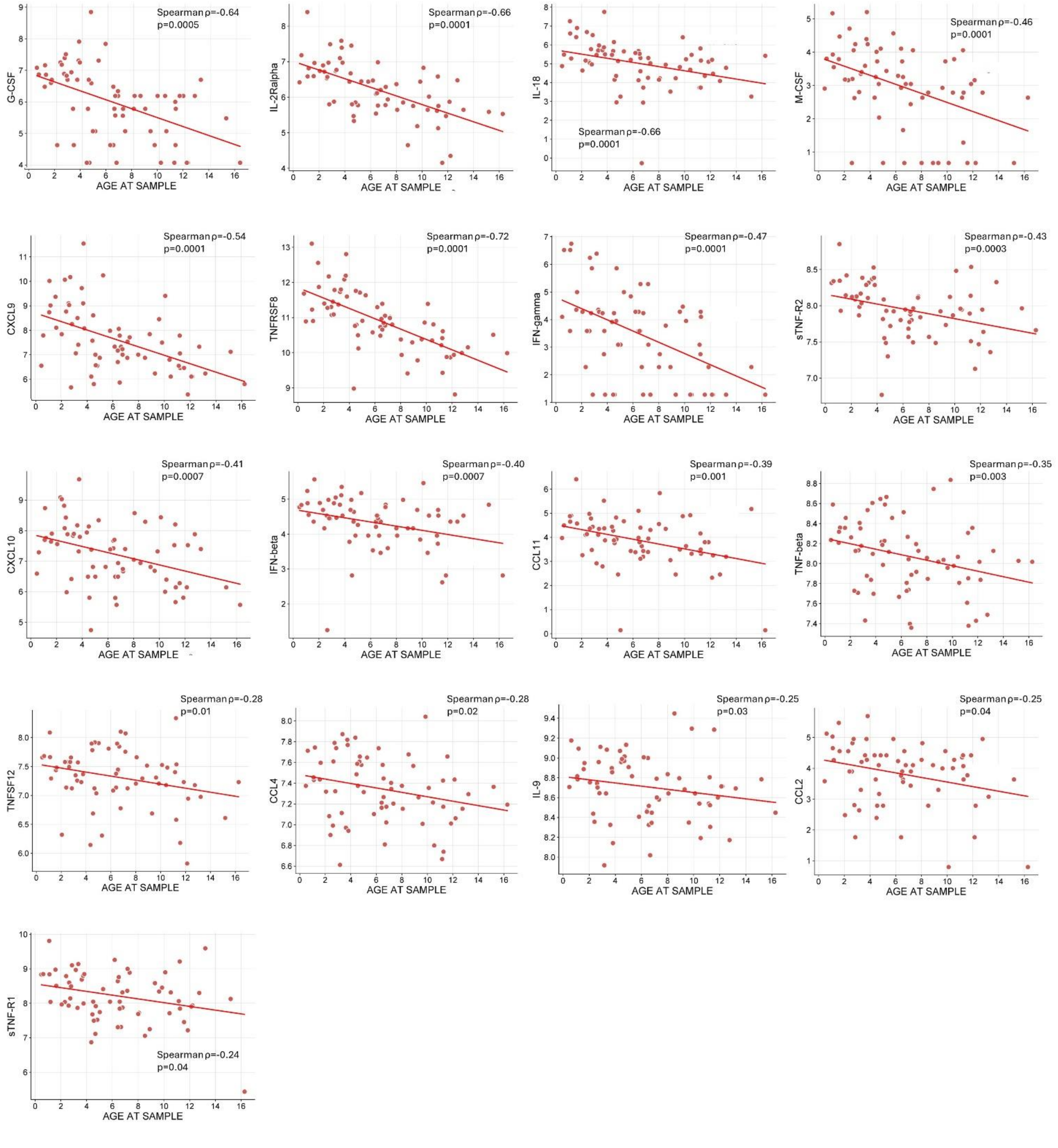


**Supplementary Figure 2.** Spearman correlation plots for each significant analyte in plasma. The plots depict age in years on the x-axis, and the concentration (log2 pg/mL) of each analyte on the y-axis. Spearman coefficient (ρ) and raw p-values are shown, n=66.

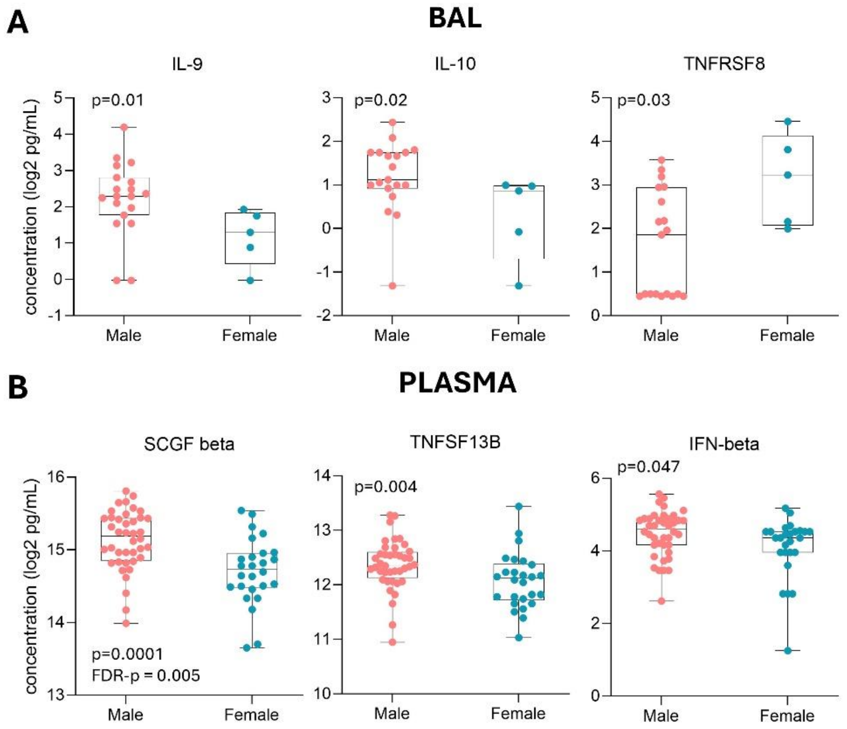


**Supplementary Figure 3.** Differences in analyte concentration between males and females in BAL and plasma. **(A)** BAL analytes that were significantly different between males and females. **(B)** Plasma analytes that were significantly different between males and females. P-values were determined by Mann Whitney U-test and p<0.05 was considered significant.

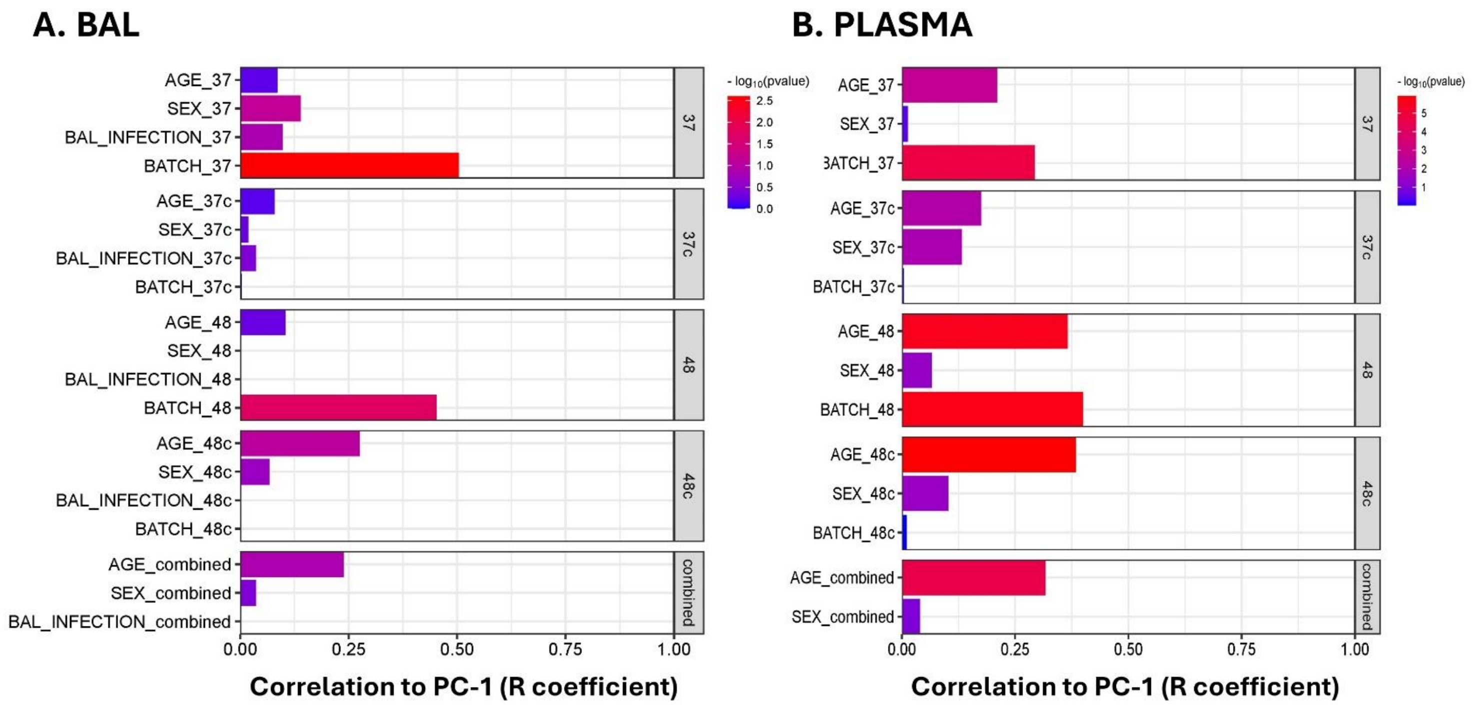


**Supplementary Figure 4**. Results of principal components analysis (PCA) on raw and ComBat batch-corrected analyte data. **(A)** PCA of BAL analyte data including age, sex, infection, and batch as variables. The first box includes uncorrected data for the 37-plex kit, the second box includes ComBat batch-corrected data for the 37-plex kit, the third box includes uncorrected data for the 48-plex kit, and the fourth box includes ComBat batch-corrected data for the 48-plex kit. The results show the removal of batch as a contributor to variation in the first dimension following ComBat correction for both kits. The fifth box shows ComBat corrected data from a combined file containing analytes of both the 37- and 48-plex kits, the results of which are presented in Figure 2 of the manuscript. **(B)** PCA of Plasma analyte data including age, sex, and batch as variables. The first box includes uncorrected data for the 37-plex kit, the second box includes ComBat batch-corrected data for the 37-plex kit, the third box includes uncorrected data for the 48-plex kit, and the fourth box includes ComBat batch-corrected data for the 48-plex kit. The results show the removal of batch as a contributor to variation in the first dimension following ComBat correction for both kits. The fifth box shows ComBat corrected data from a combined file containing analytes of both the 37- and 48-plex kits, the results of which are presented in Figure 2 of the manuscript.
